# Phenotypic and transcriptomic characterisation of a novel biallelic *RNU2-2* developmental and epileptic encephalopathy

**DOI:** 10.64898/2026.02.19.26345867

**Authors:** Olivia J. Henry, Nadja Pekkola Pacheco, Irene Duba, Magnus Burstedt, Daniel Carlberg, Angelica M. Delgado-Vega, Anna Hammarsjö, Sofie Ivarsson, Tord Jonson, Kristina Karrman, Nicole Lesko, Åsa Lindfors, Daniel Nilsson, Mia Olsson Engman, Lucía Peña-Pérez, Erik Stenund, Fulya Taylan, Malin Ueberschär, Samuel Wiafe, Sofia Ygberg, Anna Lindstrand, Anna Wedell, Ann Nordgren, Tommy Stödberg

**Author notes:** Correspondence to: Olivia J. Henry, Molecular Medicine and Surgery, Karolinska Institute, Stockholm 171 77, Sweden.

## Abstract

A significant proportion of individuals with suspected genetic developmental and epileptic encephalopathies (DEEs) remain unsolved following whole genome sequencing (WGS).

We screened individuals who received WGS analyses at Genomic Medicine Centre Karolinska for Rare Diseases for biallelic *RNU2-2* variants. Deep phenotyping was performed and phenotypic traits were transcribed to their corresponding Human Phenotype Ontology (HPO) term. HPO terms were used to generate pairwise phenotypic similarity scores and assess for significant phenotypic enrichment in the *RNU2-2* sub-cohort. RNA sequencing analyses were performed in fibroblast and blood tissues to compare splicing events between *RNU2-2* individuals and two independent control groups.

We identified 14 individuals with 12 ultra-rare biallelic *RNU2-2* variants clustering in the conserved 5’ domains. All individuals presented with a highly concordant, severe DEE, characterised by severe to profound intellectual disability, inability to walk or communicate, hyperkinesia and refractory seizures. Infantile spasms and tonic seizures were the predominant seizure types and a Lennox-Gastaut syndrome-like phenotype was common. These individuals had a significantly similar phenotypic signature when compared with 703 individuals with paediatric epilepsy (two-sided Monte Carlo permutation test, *p*=0.005). RNA sequencing analyses in fibroblast tissues showed a clear separation of aberrant mutually exclusive exon and alternate 3’ splice site events between *RNU2-2* individuals and controls, which was not detectable in blood.

In summary, we present deep phenotyping data and transcriptomic analyses which provide support for rare, 5’ clustering biallelic *RNU2-2* variants causing a novel, severe DEE. We propose an RNA sequencing methodology on fibroblast tissue for future validation of *RNU2-2* variants.

## Introduction

Developmental and epileptic encephalopathies (DEEs) represent the most severe group of epileptic disorders and present with heterogeneous aetiologies. Although a significant proportion of DEEs have a genetic basis, approximately 50% of suspected genetic DEEs remain unsolved following whole exome or whole genome sequencing (WGS)^1^.

Advances in genomic technologies have facilitated the identification of novel pathophysiological mechanisms in DEEs including poison exon inclusion, oligogenic inheritance and brain somatic mosaicism^2^. Recently, dominant variants in non-coding small nuclear RNA (snRNA) genes *RNU2-2*^3–5^, *RNU4-2*^6,7^ and *RNU5B-1*^4,8^ have been identified as novel causes of neurodevelopmental disorders (NDD) and DEEs, with pathogenic heterozygous variants in *RNU2-2* characterised by a phenotype with prominent epilepsy.

*RNU2-2* codes for the U2 snRNA which is a central component of the major spliceosome. Correct splicing requires a concerted series of dynamic compositional and structural changes in the spliceosomal small nuclear ribonucleoproteins (snRNPs) and associated proteins to generate functional mature mRNA from the more than 99% of pre-mRNA that is spliced by the major spliceosome^9^. Dysregulated splicing has been implicated in multiple genetic diseases, including the minor spliceosomopathies caused by biallelic variants in *RNU4ATAC* (MIM 601428) and *RNU12* (MIM 620204), in which intron retention has been shown to underlie the pathophysiological mechanism^10^.

Since 2015, we have accrued a large clinically ascertained cohort of individuals with epilepsy and NDD who have received clinical WGS at the Genomic Medicine Centre Karolinska for Rare Diseases (GMCK-RD)^1,11,12^. In line with three recent preprints^13–15^, we describe a cohort of 14 individuals with a novel recessive *RNU2-2* disorder harbouring ultra-rare biallelic *RNU2-2* variants who share a highly concordant, severe DEE phenotype. Despite the relatively high frequency of individuals presenting with this disorder at GMCK-RD, these variants were previously missed on prior comprehensive research genetic investigations, reflecting challenges in prioritising and classifying non-coding variants using standard pipelines. Indeed, the large-scale population database at Genomics England 100,000 Genomes Project was necessary for the identification of the statistically significant, recurrent, and enriched *de novo* variants in *RNU2-2* and *RNU4-2*^3,4,6,7^. This approach is not currently feasible in a clinical setting and difficult to validate with novel/non-recurrent recessive variants, highlighting the need for novel tests to validate such variants. In this study, we demonstrate strong support for a distinct recessive *RNU2-*2 disorder using comprehensive phenotyping, Human Phenotype Ontology^16^ (HPO)-based similarity analysis and RNA sequencing analyses across blood and fibroblast tissues, with the latter test providing a potential novel clinical validation approach for *RNU2-2* variants.

## Methods

### WGS analytical workflow at GMCK-RD

All individuals were investigated with clinical WGS through the GMCK-RD (Karolinska University Hospital). GMCK-RD is a multidisciplinary, academic-clinical partnership that promotes a collaborative workflow^11,12^. Individuals with suspected rare genetic disorders are referred by their primary clinicians for WGS through the Centre for Inherited Metabolic Diseases (CMMS) and/or the Department of Clinical Genetics and Genomics (KGG) at GMCK-RD.

The CMMS workflow integrates expertise across clinical genetics, paediatrics, neurology, endocrinology, molecular genetics, bioinformatics and biochemistry to provide genetic diagnostics for individuals with suspected genetic metabolic diseases and complex epilepsies^1,11^. The KGG department receives referrals from patients presenting with a broad range of disorders including neurodevelopmental disorders and malformation syndromes.

In addition to regionally referred clinical cases, the GMCK-RD framework also includes cases referred through national and international research collaborations, including the Undiagnosed Diseases Network Sweden (UDN Sweden) and the Karolinska Undiagnosed Diseases Program, which is part of the Undiagnosed Diseases Network International (UDNI)^17^.

WGS was performed on all patients and parents according to clinical protocols and processed using our in-house developed Mutation Identification Pipeline (MIP)^11^. MIP incorporates a local database called loqus database (loqusdb) which annotates the frequency of all WGS variants at GMCK-RD at the family level^18^. Clinical analyses are limited to disease-associated genes, typically via panel-based filtering. At both CMMS and KGG, physicians assess patient phenotypic data to select appropriate *in-silico* gene panels (e.g. epilepsy panel or intellectual disability panel) or generate customisable HPO or PanelApp gene panels. If clinical analyses fail to identify a causative variant(s), and the individual/parents/guardians have provided informed consent, the WGS data may be investigated further through a research track where clinical filters are removed and all genes are analysed.

### RNU2-2 biallelic variant screening

Following release of the dominant *RNU2-2* disorder preprint^19^ we searched for *RNU2-2* variants in individuals in loqusdb who had previously undergone research-based analyses, identifying eight individuals from five families with biallelic variants and strikingly compatible phenotypes. Following the final publication of the dominant *RNU2-2* disorder^3,4^ we screened all families/singletons from CMMS and KGG in loqusdb for *RNU2-2* variants up until September 9^th^, 2025. Variants were initially prioritised based on zygosity, population frequency (gnomAD v4.1.0 non-UK biobank (UKB) variant frequencies ≤2 homozygotes), position (within the snRNA-encoding region) and phenotypic concordance. After reviewing data from the initial screening, variants were further filtered to exclude variants present in homozygous state in gnomADv4.1.0 non-UKB or in non-NDD from the 100,000 Genomes Project^14^ and/or lacking at least one variant at n.61 or earlier in *RNU2-2* where variation is depleted in large population datasets^4^, consistent with constraint. In addition to our study, the biallelic *RNU2-2* disorder was recently independently described in three preprints^13–15^. Subsequently, variant level data and broad phenotypic information (yes/no for epilepsy, intellectual disability and brain MRI abnormalities) from 13 individuals from the current cohort will be included in another publication^14^. All families included in the current study provided informed consent. Ethical approval was obtained from the Swedish ethical review board (Dnr 2022-06033-01, Dnr 2008/351-31).

### Phenotyping in individuals harbouring biallelic RNU2-2 variants

Medical records were reviewed for each patient. Prior to receiving WGS patients undergo comprehensive investigations including brain MRI, metabolic work up, etc, to exclude other plausible aetiologies. Where applicable phenotypic traits were transcribed to their corresponding HPO term. Seizure classifications and syndrome diagnoses were allocated according to the ILAE classifications^20–22^. Individuals allocated Lennox-Gastaut syndrome (LGS)-like fulfilled all criteria for LGS except one or both of the mandatory EEG criteria^22^.

### HPO similarity analyses

The epilepsy cohort utilised for phenotype similarity analyses included 703 individuals with paediatric epilepsy who underwent WGS at CMMS between 2014-2022 and has been described in detail previously^1^. All those individuals were screened for a specific subset of 16 HPO terms (Supplemetary data) to ensure a consistent depth of information. HPO terms for the *RNU2-2* individuals were transcribed to their broader corresponding HPO term where applicable to prevent bias (for example, severe intellectual disability (HP:0010864) was represented as intellectual disability (HP:0001249)). Analyses were limited to one individual per family in both the *RNU2-2* cohort and the broader epilepsy cohort.

Phenotypic analyses were performed in R using the OntologyIndex and OntologySimilarity packages^23^. Pairwise phenotypic similarity scores were generated using the OntologySimilarity package. Redundant or duplicated HPO terms per individual were removed. The *RNU2-2* cohort (*n*=9 with siblings excluded) phenotypic similarity scores were compared with the broader epilepsy cohort (excluding siblings and the *RNU2-2* individuals, *n*=703) using a two-sided Monte Carlo permutation test. A null distribution was generated by recomputing the phenotypic similarity score for 1000 randomly sampled groups of nine individuals from the broader epilepsy cohort, and an empirical *p*-value was calculated based on the extremeness of the observed score. HPO term enrichment analyses were performed using Fisher’s exact test with the Benjamini-Hochberg *p*-value adjustment to control for multiple testing.

### Variant confirmation

Sanger sequencing was performed on genomic DNA from all probands, their parents and two unaffected siblings from family 6 under standard conditions to confirm the presence of each *RNU2-2* variant, verify the trans configuration in parents and segregate across unaffected siblings. Primers RNU2-2_F:CGGGTTTATGCAAAACAAACTG, RNU2-2_R:CAACAAGACACTCAAACACGC were designed to amplify the entire ENST00000410396.1 transcript. Amplicons were sequenced using BigDye Terminator cycling chemistries (Applied Biosystems) on a 3500xl DNA sequencer (Applied Biosystems).

### RNA sequencing

#### Sequencing

Where available, blood and fibroblast samples were obtained from individuals with biallelic *RNU2-2* variants, as well as from nine individuals with recurrent pathogenic *RNU4-2* variants n.64_65insT (*n*=7), n.65A>G (*n*=1) and n.69C>T (*n*=2) to serve as a validation cohort. Samples were prepared according to the same protocol at CMMS and sequenced at Clinical Genomics. Total RNA was isolated from whole blood and cultured fibroblasts using PAXgene RNA blood kit (Qiagen) and RNeasy mini kit (Qiagen), respectively. Generation of RNA sequencing libraries was performed using the Illumina Stranded mRNA library preparation kit/Illumina TruSeq Stranded mRNA following the manufacturer’s instructions. mRNA from blood and fibroblasts was extracted using poly (A) capture with magnetic beads and poly (T) oligos, then sheared and reverse-transcribed. Paired-end 150bp sequencing was performed on NovaSeq 6000 (Illumina). Conversion Software bcl2fastq v2.20 (Illumina) was used to demultiplex the samples, which were then processed using our in-house bioinformatics pipeline Tomte (https://github.com/genomic-medicine-sweden/tomte) and further downstream analysis.

#### RNA-seq data processing and analysis

Outlier analysis of gene expression and splicing was performed using the Detection of RNA Outliers Pipeline (DROP)^24^ within Tomte, using internal control datasets for each tissue type (*n=*195 control samples for blood, *n*=64 control samples for fibroblast). Outlier events were defined as having *p*<0.05, and results were further filtered by relevant gene lists.

Differential splicing analysis was performed using rMATS-turbo (v4.3.0)^25^ with default parameters. Bam files from blood samples were first filtered to remove 99.9% of haemoglobin reads then all bam files were downsampled to 120 million read pairs. Two groups of age- and sex-matched controls were used for independent differential analyses. rMATS results were filtered for coverage >10 reads and Percent Spliced In (PSI) values between 0.05 and 0.95. Significant events were further defined as false discovery rate <0.1 and deltaPSI > 0.05. Principle component analysis was performed on these event sets, including a validation step to plot the second control set by significant events found between the first control set and patients. Filtering and some plotting of rMATS results was adapted from https://github.com/Xinglab/rmats-turbo-tutorial.

Differential expression analysis was performed using DESeq2 (v1.48.2) using salmon quantification files from the Tomte pipeline. The same control cohorts were used in differential splicing and expression analyses. Genes with ≥10 total counts were used for DESeq2 analysis, and *p*-adjusted < 0.05 and |log2 fold-change| >1 was considered significant.

### Statistical analysis

Model fit and assumptions were evaluated using standard diagnostics appropriate to each analysis type. For permutation-based similarity scoring, the null distribution was inspected for symmetry and stability across 1,000 permutations. For differential splicing, PSI distributions and coverage were examined to ensure reliable quantification. For DESeq2 differential expression, dispersion estimates and residuals were inspected and p-value distributions were assessed for conformity to null expectations. Multiple testing correction was applied as described in the relevant methods subsections.

## Results

### Ultra-rare biallelic RNU2-2 variants segregate with a DEE phenotype

28,196 singletons/families (3,790 from CMMS and 24,406 from KGG, including 169 from UDN Sweden) were screened for rare biallelic *RNU2-2* variants. Fourteen individuals were identified across nine families who harboured 12 ultra-rare (MAF≤0.0000723 in gnomADv4.1.0 non-UKB) biallelic *RNU2-2* variants (table1) and shared compatible DEE phenotypes (table 2).

All individuals had previously undergone multiple clinical WGS analyses (including in 2025), and most had also undergone research analyses (11/14) without identification of other plausible candidates. Most variants (10/12) occurred in compound heterozygous states except in three individuals from two families who harboured homozygous variants (n.61C>G and n.23A>G). All variants were confirmed to be in trans and segregated with disease, with each unaffected parent harbouring a single variant, and two healthy siblings in family 1 harbouring only a single variant each. All variants except n.14C>G were recurrent either in our cohort or were reported in affected individuals in other biallelic *RNU2-2* preprints with the same base change or at the same position (table 1, figure 1)^13–15^. All individuals harboured at least one variant located in one of the conserved 5’ domains (figure 1) at position n.61 or earlier. Only 3/12 variants were located outside the 5’ region: n.104T>G in the conserved Sm binding region, n.107_118del close to the Sm binding region and n.131G>Cin stem loop III. The variants are depleted in both gnomAD v4.1.0 non-UKB and non-biallelic *RNU2-2* families at CMMS and KGG at pathogenic variant regions (figure 2a). All pathogenic variants were conserved, with most occurring at peak PhyloP scores from the Zoononia 241-mammal alignment (figure 2b). Two individuals harboured biallelic *RNU2-2* variants of interest however were excluded due to incompatible phenotypes and either the second variant in trans being more common (73 heterozygotes in gnomAD v4.1.0 non-UKB) or harbouring a causative pathogenic variant in another gene (supplementary data). No other individuals within the dataset harboured two rare variants in regions of interest.

**Figure 1.**
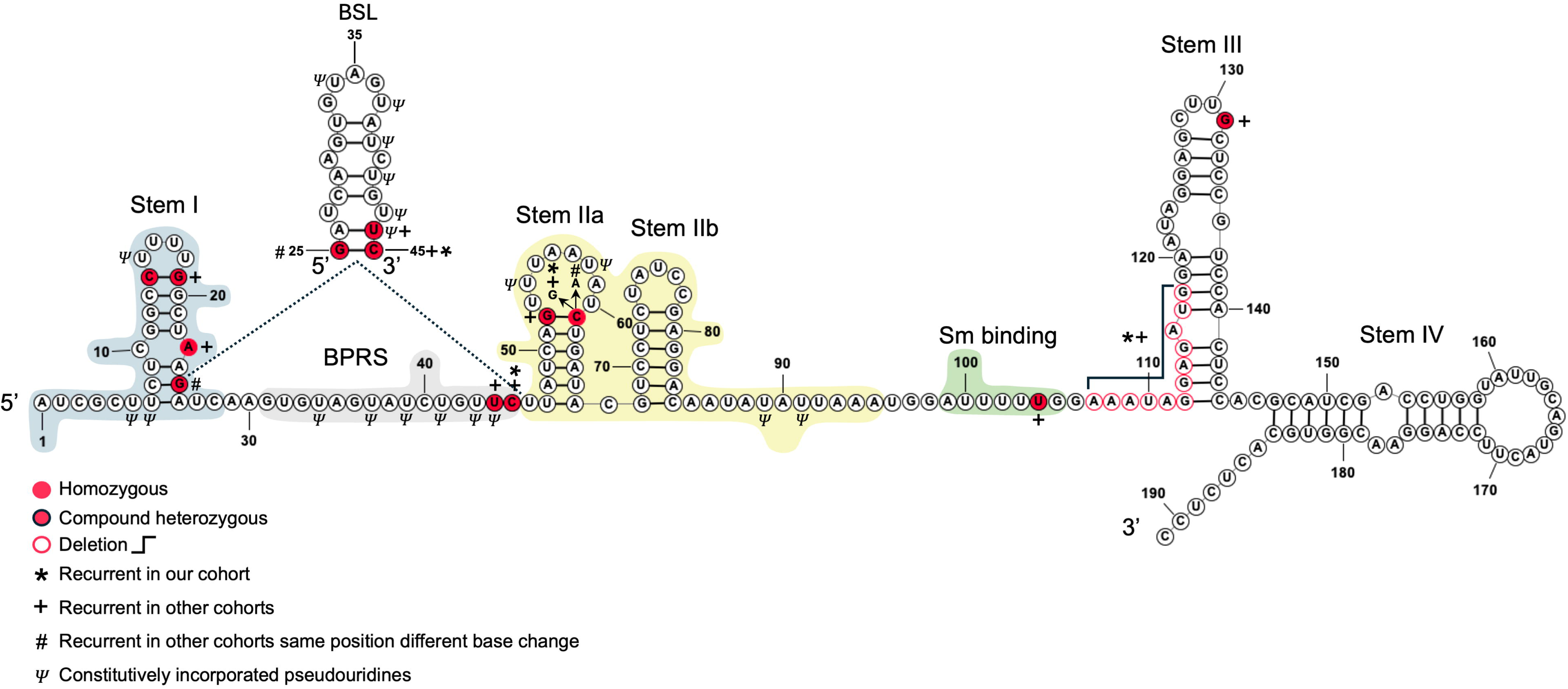
RNU2-2 secondary structure^27^ with causative variants identified in the cohort highlighted. Conserved functional domains Stem I, BPRS, Stem II and Sm binding are highlighted in blue, grey, yellow and green, respectively. Note: n.61 contains one homozygous n.61C>G individual and compound heterozygous n.61C>G and n.61C>A variants in *n*= 2 and *n*=1 families, respectively. The BSL (17S U2 snRNP) forms early in spliceosome assembly before unwinding to base pair with the intronic splice site.

**Figure 2.**
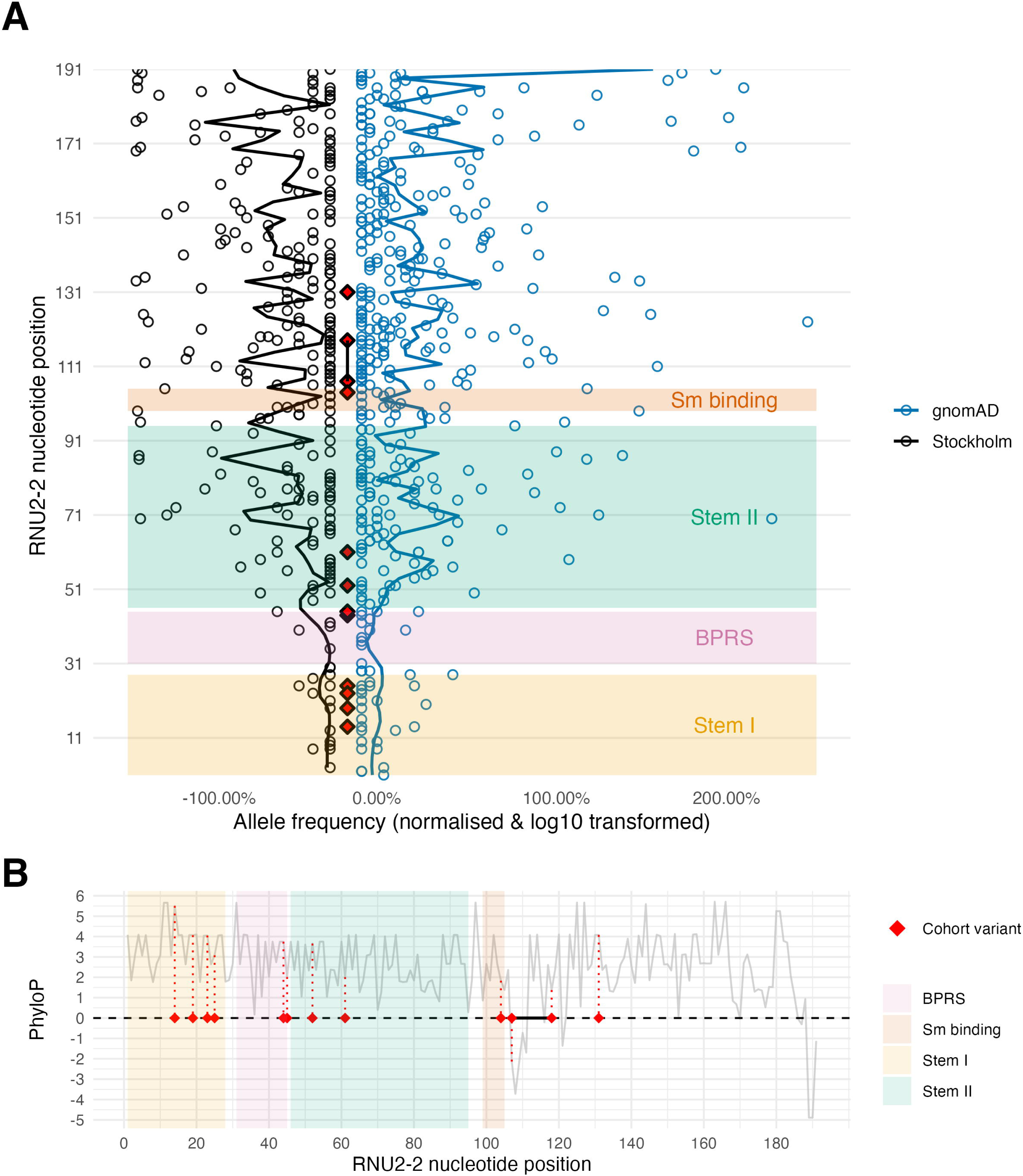
*RNU2-2* cohort variants conservation and frequency in population datasets. **A)** Conservation as measured by PhyloP scores from the Zoononia 241-mammal alignment. The two diamonds connected by a solid black line indicate the n.107_118del. All other variants are SNVs. Conserved functional regions are highlighted. **B)** *RNU2-2* heterozygous variant frequencies across individuals who received WGS at CMMS and Clinical Genetics and Genomics at Karolinska University Hospital in Stockholm (black) compared with the corresponding variant frequencies in gnomAD (v4.1.0 non-UKB) (blue). Frequencies are normalised to the total number of individuals who received WGS across each cohort and log10 transformed for visualisation purposes. The smoothed line applies a locally weighted regression trend line from the nearest 3% of data points. Red diamonds alone denote the positions of biallelic *RNU2-2* patient SNVs and red diamonds connected by a line denote a deletion spanning both diamonds. Conserved functional regions are highlighted. For smaller insertions or deletions only the starting position of these variant types is graphed.

**Table 1.**
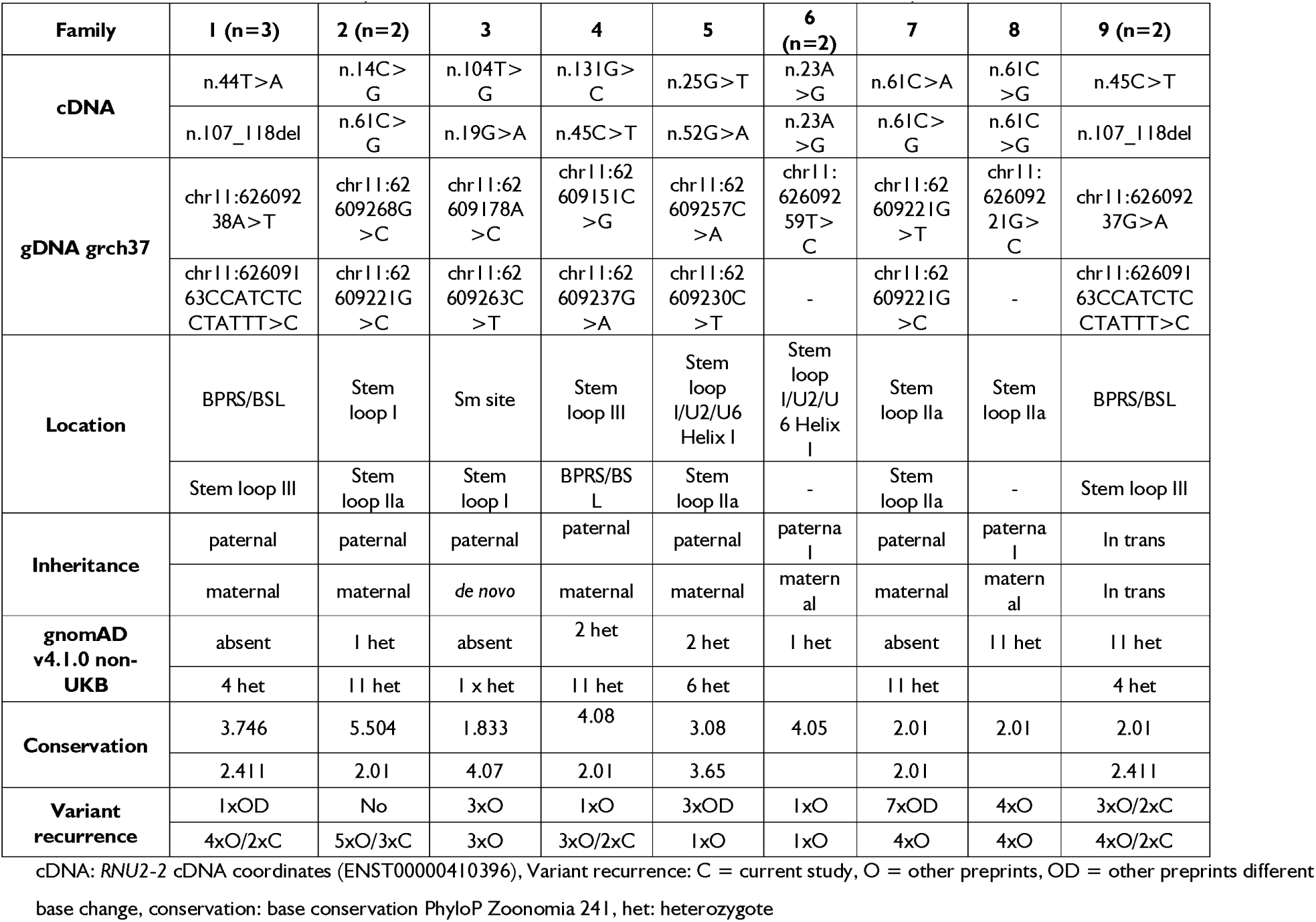
*RNU2*-2 variants in each family. Total affected individuals are shown in brackets, otherwise only one individual is affected.

**Table 2.** Deep phenotyping data from individuals with biallelic *RNU2*-2 variants.

| Individual | 1A | 1B | 1C | 2A | 2B | 3 | 4 | 5 | 6A | 6B | 7 | 8 | 9A | 9B |
| --- | --- | --- | --- | --- | --- | --- | --- | --- | --- | --- | --- | --- | --- | --- |
| Gender | male | female | female | male | male | male | male | male | male | female | female | female | male | male |
| Developmental stage & seizure type at onset | Neonatal tonic | Infancy IS | Infancy IS | Childhood FIC motor | Infancy IS | Infancy IS | Infancy GA | Infancy IS | Infancy IS | Infancy FIC | Infancy IS | Childhood BTCS | Infancy IS | Infancy IS |
| Seizure types | GT, GTC, FIC motor, FIC, GC | IS, GT, GTC, AS, focal motor | IS, ES, GT, EMA, GA, GM | FIC motor, ES, GT | IS, GT, possible ES | IS, ES, tonic, possible GM | GA, GTC/BTC, GM, FIC, FIC motor, FPC motor, focal tonic | IS, ES, GT, AA, GA, GTC, BTC | IS, GMA, FIC tonic, FIC, FIC clonic | IS, GT, AA, FIC motor, FIC tonic, BTCS | IS, ES, tonic, AS, GTC | BTC, GA, GM, FIC, focal tonic | IS, ES, GTC, GT, GM, GA | IS |
| Epilepsy Syndrome | LGS-like | IESS, LGS-like | IESS, LGS-like | LGS-like | IESS, LGS-like | IESS, LGS | DEE | IESS, LGS | IESS, LGS | IESS, LGS | IESS, LGS | N | IESS, LGS | IESS, LGS |
| Refractory seizures | Y daily | Y daily | Y daily | Y weekly | Y daily | Y daily | Y daily | Y periods of daily seizures | Y daily | Y daily | Y daily | Y periods of daily seizures | Y | Y |
| GDD | Y | Y | Y | Y | Y | Y | Y | Y | Y | Y | Y | Y | Y | Y |
| ID | Profound | Profound | Profound | Severe | Severe | Profound | Severe | Profound | Profound | Severe | Severe | Severe | Severe | Severe |
| Regression | Y | Y | Y | Y | Y | Y | Y | Y | Y | Y | Y | Y | Y | Y |
| Absent speech | Y | Y | Y | Y | Y | Y | Y | Y | Y | Y | Y | Y | Y | Y |
| Non ambulatory | Y | Y | Y | Y | Y | Y | Y | Y | Y | Y | Y | Y | Y | Y |
| Involuntary movements | Choreo, dystonia | Choreo, dystonia | Choreo, myoclonus, dystonia | Choreo, hyperkinesia, dystonia | Choreo | Choreo | Hyperkinesia, dystonia | N | Choreo | N | Myoclonus | Dystonia & involuntary movements | Myoclonus | Myoclonus |
| Motor stereotypy | NA | N | NA | Y | Y | N | Y | Y | Y | Y | Y | Y | Y | N |
| Voluntary movement | N | N | N | Y | Y | N | Y | N | Y | Y | Y | Y | Y | N |
| Abnormal tone | Y | Y | Y | N | Y | Y | Y | Y | Y | Y | Y | Y | Y | Y |
| Interactions | N | Very limited | Y | Very limited | Very limited | N | Y | Very limited | Y | Very limited | N | Y | Very limited | N |
| Microcephaly | Y | Y | Y | N | N | N | N | Y | N | N | Y | N | N | N |
| Brain atrophy | Y | Y | Y | Y progressive | Y | Y | N | Y progressive | N | N | N | Y progressive | Y | Y |
| Tube feeding | Y | Y | Y | Y | N | Y | Y | Y | N | N | Y | Y | N | N |
| Bruxism | Y | N | N | Y | Y | N | N | Y | N | N | N | Y | Y | Y |

Y: Yes, N: No, y: years, m: months, ID: intellectual disability, regression: developmental regression, GDD: global developmental delay choreo: choreoathetosis, myoclonus: non-epileptic myoclonus, AA: generalised atypical absence seizure, AS: absence seizure, BTC: bilateral tonic clonic seizure, EMA: eyelid myoclonia with absence, ES: epileptic spasms, IESS: infantile epileptic spasms syndrome, IS: infantile spasms, FIC: focal impaired consciousness seizure, focal motor: focal seizure with motor manifestations, FPC: focal preserved consciousness seizure, GA: generalised atonic seizure, GC: generalised clonic seizure, GT: generalised tonic
seizure, GM: generalised myoclonic seizure, GTC: generalised tonic clonic seizure, LGS: Lennox-Gastaut syndrome, motor: with motor manifestations. Neonatal period: 0 - 28 days, infancy: 29 days – 24 months, childhood: 25 months – 18 years.

### The biallelic *RNU2-2* disorder is highly prevalent in paediatric epilepsies

Individuals with suspected genetic epilepsies are largely referred to CMMS for WGS analysis at GMCK-RD. The frequency of families with biallelic pathogenic *RNU2-2* variants within this sub-cohort was 0.65% (7/1075 families). The overall frequency at CMMS was 0.18% (7/3790), 0.02% (5/24406) at KGG and 0.032% (9/28196) across all families/singletons at CMMS and KGG.

### Individuals with biallelic *RNU2-2* disorder present with a highly concordant phenotype

#### Phenotypic similarity analyses

HPO-based phenotypic similarity analyses comparing one individual from each family with biallelic *RNU2-2* (*n*=9 individuals) with the broader epilepsy cohort (*n*=703 individuals) demonstrate that the biallelic *RNU2-2* patients had a significantly similar phenotypic signature (similarity score 6.361; two-sided Monte Carlo permutation test, *p*=0.005,) when compared with the broader epilepsy cohort (permutation mean similarity score 5.327, standard deviation 0.343, figure 3). HPO terms which were significant enriched in biallelic *RNU2-2* individuals are summarised in table 3 and figure 3b.

**Figure 3.**
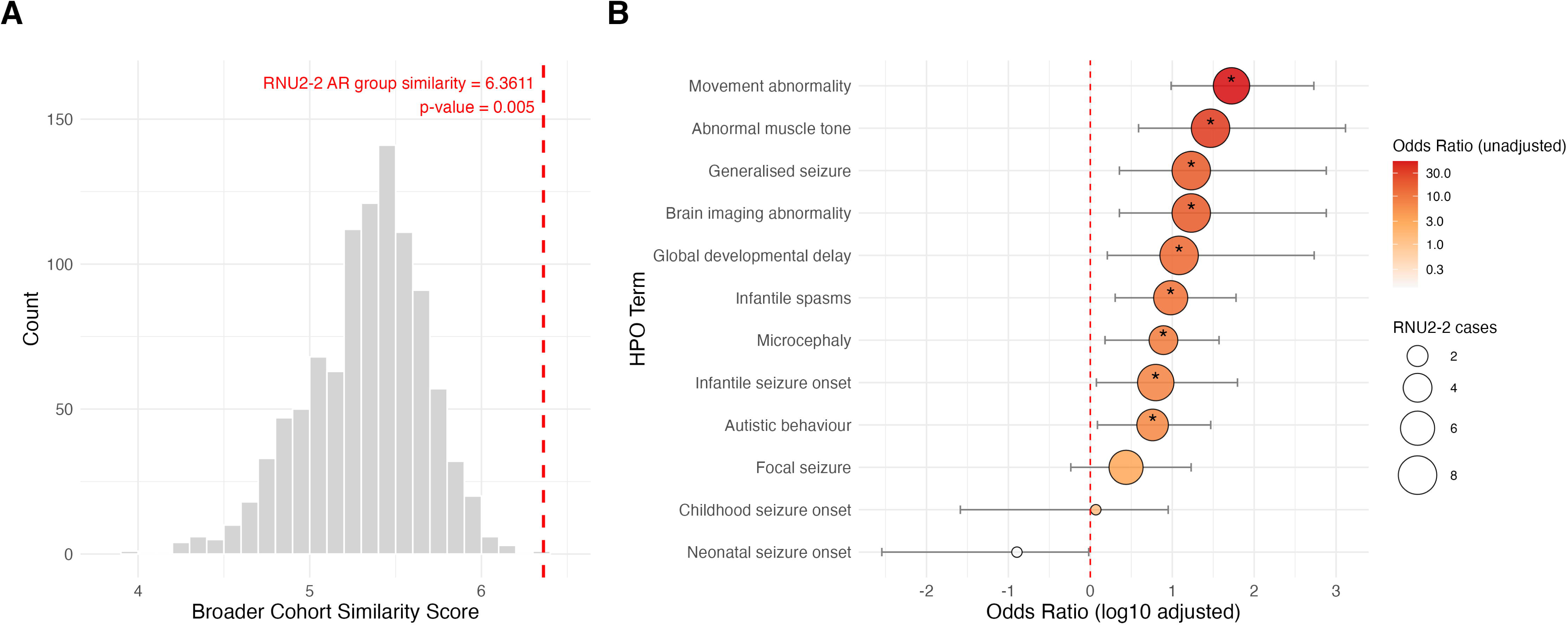
Phenotype similarity analyses. **A**) Broader epilepsy cohort similarity scores based on 1000 permutations compared with the *RNU2-2* cohort. Grey bars denote permutations, dashed red line illustrated *RNU2-2* individuals similarity score. **B)** HPO term enrichment in the *RNU2-2* cohort. Significantly enriched terms (p≤0.05) are denoted with stars. Intellectual disability was excluded due to a high odds ratio for visualisation purposes.

**Table 3.**
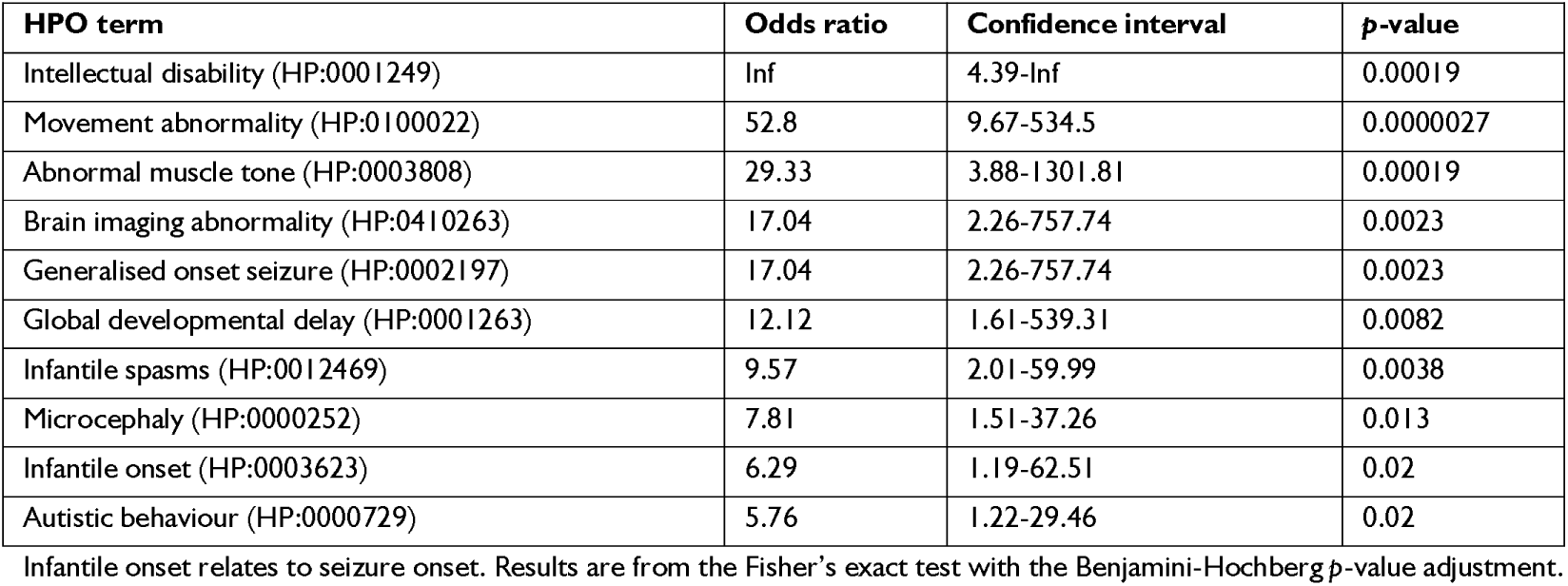
HPO terms significantly enriched in biallelic *RNU2*-2 individuals compared with the broader epilepsy cohort.

| HPO term | Odds ratio | Confidence interval | p-value |
| --- | --- | --- | --- |
| Intellectual disability (HP:0001249) | Inf | 4.39-Inf | 0.00019 |
| Movement abnormality (HP:0100022) | 52.8 | 9.67-534.5 | 0.0000027 |
| Abnormal muscle tone (HP:0003808) | 29.33 | 3.88-1301.81 | 0.00019 |
| Brain imaging abnormality (HP:0410263) | 17.04 | 2.26-757.74 | 0.0023 |
| Generalised onset seizure (HP:0002197) | 17.04 | 2.26-757.74 | 0.0023 |
| Global developmental delay (HP:0001263) | 12.12 | 1.61-539.31 | 0.0082 |
| Infantile spasms (HP:0012469) | 9.57 | 2.01-59.99 | 0.0038 |
| Microcephaly (HP:0000252) | 7.81 | 1.51-37.26 | 0.013 |
| Infantile onset (HP:0003623) | 6.29 | 1.19-62.51 | 0.02 |
| Autistic behaviour (HP:0000729) | 5.76 | 1.22-29.46 | 0.02 |

Infantile onset relates to seizure onset. Results are from the Fisher's exact test with the Benjamini-Hochberg *p*-value adjustment.

#### Deep phenotyping

Most individuals had been under genetic investigation for many years at GMCK-RD with a median referral age of 55 months (range 6 months to19 years). The median time from referral to a last follow up was 8 years and 3 months (range 1 month to 14 years 3 months) at a median age of 12 years 6 months (range 5 years and 6 months to 28 years and 3 months). Nine of the fourteen individuals were male. Phenotypic data are summarised in figure 4ab, table 2 and supplementary table 1. Global developmental delay was evident in all individuals, presenting at a median age of 5.5 months (range birth to 12 months). Unsupported sitting was achieved in 9/14 individuals (64%), however many were delayed (median age 9.5 months, range 5 to 31 months). Crawling was achieved by only three patients at 26 months or later. Independent walking was achieved in only one individual at 4.5 years however she soon after lost this skill with seizure onset. All patients had severe (8/14, 57%) to profound (6/14, 43%) intellectual disability and were non-verbal with severely limited/absent social interactions. Developmental regression was observed in all individuals. All individuals had severely limited motor capabilities, with nearly half of all individuals (6/14, 43%) without any voluntary movement. Hypotonia and hypertonia was present in 11/14 (79%) and 11/14 (79%) of individuals, respectively, with most individuals presenting with both. Movement abnormalities were seen in most individuals (12/14, 86%) and presented as dystonia, choreoathetosis, hyperkinesia or non-epileptic myoclonus. Most individuals (10/14, 71%) showed atrophy on brain MRI, with only two individuals without any abnormalities detected. Other prevalent features included tube feeding (9/14, 64%), bruxism (7/14, 50%), short stature (7/14, 50%), recurrent/severe infections (11/14, 79%) and sleep disturbances (11/14, 79%). No obvious facial features were noted overall; however, some patients presented with long palpebral fissures, and features consistent with progressive orofacial hypotonia were observed.

**Figure 4.**
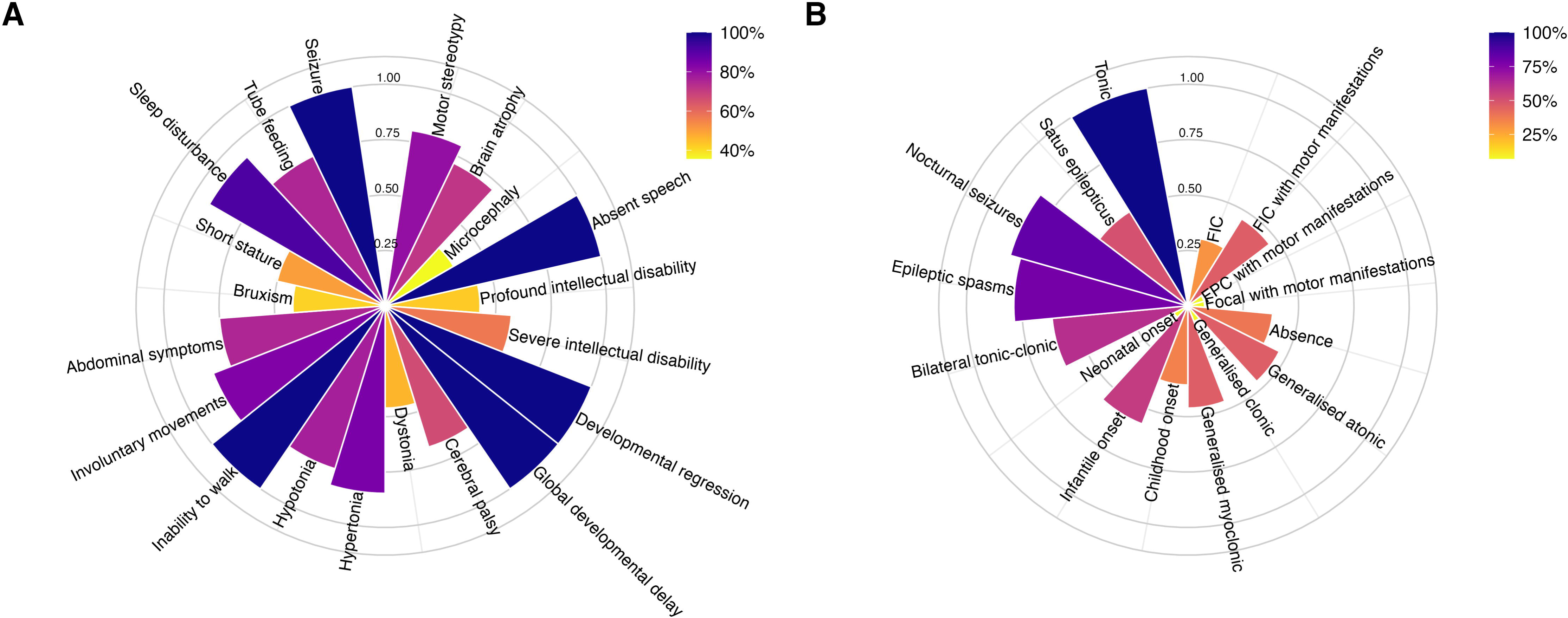
Deep phenotyping HPO traits in the *RNU2-2* cohort. **A)** Comorbidities (non-seizure related). **B)** Seizure types and traits.

All individuals had refractory seizures which commenced early in life (onset ≤2 years in 12/14 individuals, median onset age 10.5 months) and persisted until last review. The most prevalent seizure types were tonic seizures (13/13, 100%), epileptic spasms (11/14, 79%) and bilateral tonic-clonic seizures (8/13, 62%). While generalised seizures were significantly enriched in our cohort, 7/13 individuals (54%) also had focal seizures. More than half the cohort had experienced status epilepticus (8/14, 57%) and most individuals (12/14, 86%) had nocturnal as well as diurnal seizures. Most individuals (12/14, 86%) had an LGS(/-like) epilepsy syndrome diagnosis, often progressing from infantile epileptic spasms syndrome (10/14, 71%).

### Biallelic *RNU2-2* variants alter splicing in a tissue specific manner

RNA-sequencing was performed on blood and fibroblasts from individuals in the *RNU2-2* cohort (*n*=12 blood and *n*=9 fibroblasts), *RNU4-2* cohort *(n=*6 blood and *n*=5 fibroblasts), and two independent control groups (*n*=8 blood and *n*=10 fibroblasts, *n*=9 blood and *n*=10 fibroblasts). Since *RNU2-2* is required for correct pre-mRNA splicing, we assessed differential splicing using rMATS-turbo, comparing the *RNU2-2* cohort with controls across all splicing events and significant events using principle component analysis of PSI values. To serve as a positive control, we employed the same rMATS analyses on a cohort of individuals with known pathogenic *RNU4-2* variants since this methodology has previously identified splicing abnormalities associated with *RNU4-2* variants^8^.

Across all splicing events in fibroblasts there was clear separation of mutually exclusive exon and alternate 3’ splice site events between *RNU2-2* individuals and controls (figure 5a-b). Across all splicing events in blood, no differences were detected (supplementary figure 1a-e). In significant events in both fibroblasts and blood, all event types in the *RNU2-2* cohort separated from controls with a higher number of significant events occurring in fibroblasts (figure 5c-d, supplementary figure 2a-h). Significant events were validated with a second independent control group, with both control groups separating from the *RNU2-2* individuals across both blood and fibroblasts. We sought to cross validate the rMATS-turbo splicing analyses using FRASER^26^ through DROP. *RNU2-2* individuals showed a trend towards elevated aberrant splicing, which was more apparent in the CMMS epilepsy panel genes (version 22: https://www.karolinska.se/4af5ef/globalassets/global/2-funktioner/funktion-kul/cmms/genetik/ep-v22.pdf), however this did not reach statistical significance (supplementary figure 5b, 5d).

**Figure 5.**
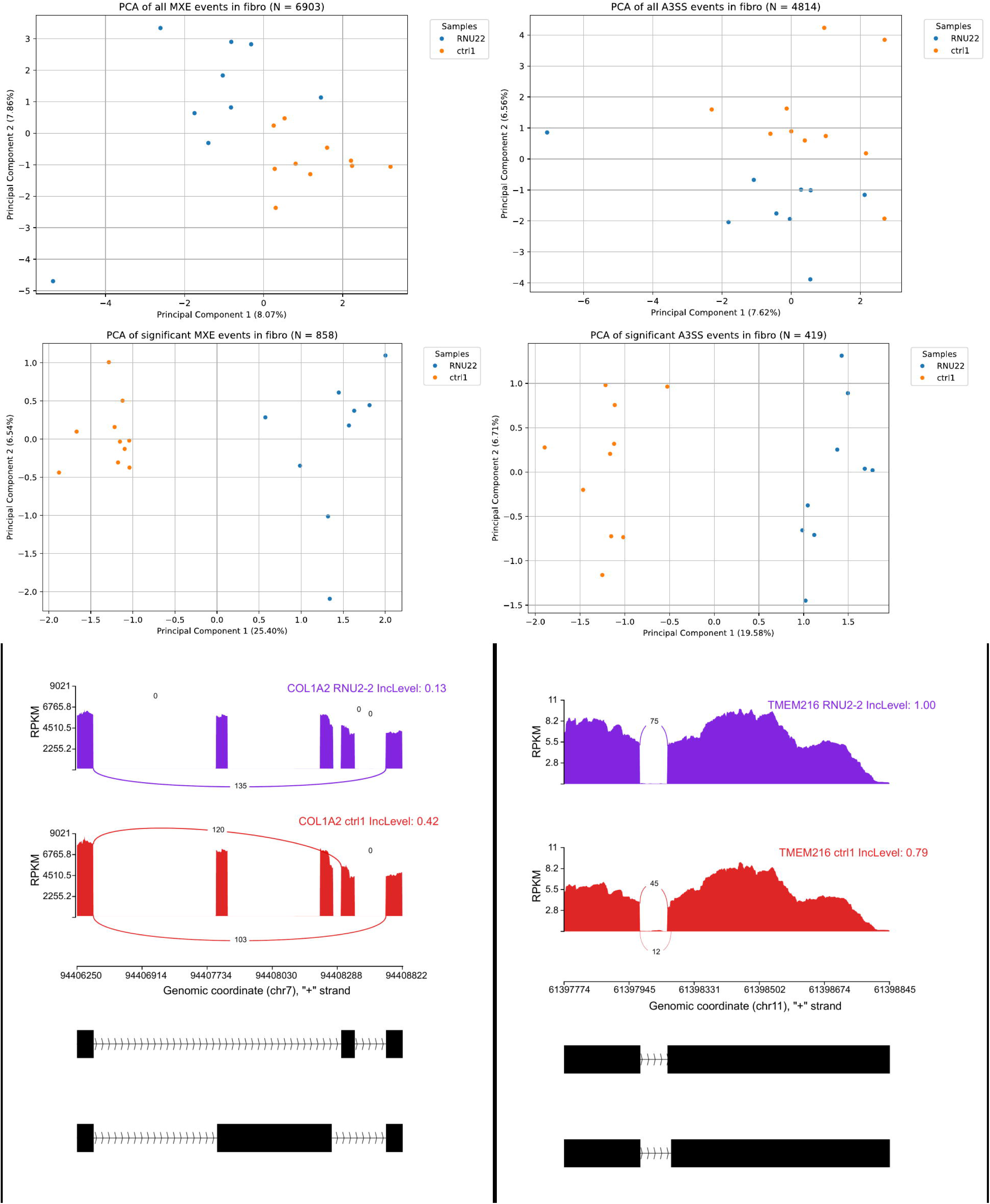
rMATS-turbo splicing analyses in the *RNU2-2* cohort compared with controls (RNA isolated from fibroblasts). **A)** Mutually exclusive exons: all events **B)** 3’ splice sites: all events. **C)** Mutually exclusive exons. significant splicing events. **D)** 3’ splice sites significant splicing events. **E)** Sashimi plot demonstrating an isoform shift in *COL1A2* due to a mutually exclusive exon event. *RNU2-2* individuals only display splicing at a non-canonical splice site (TGCCATCG/TATG). Controls display splicing at the canonical splice site (ATGGACAG/GATT) and the non-canonical splice site (TGCCATCG/TATG). **F)** Sashimi plot demonstrating an isoform shift in *TMEM216* due to an alternate 3’ splice site event.

In all splicing events in *RNU4-2* individuals, there was separation of all event types in fibroblasts and alternative 5’ splice site events in blood (supplementary figure 3a-f). Across significant events there was clear separation of all event types in the *RNU4-2* cohort in fibroblasts and blood (supplementary) which was seen across both control sets. There was a comparable number of significant events across both tissues except for mutually exclusive exons where more than twice as many significant events were detected in fibroblasts compared with blood (supplementary figure 4a-b). In cross validation analyses using FRASER *RNU4-2* individuals showed significantly more aberrant splicing events than controls in fibroblasts (supplementary figure 5b, 5d). Neither the *RNU2-2* or the *RNU4-2* cohort displayed significant aberrant expression on DROP analyses (supplementary figure 5a, 5c). DESeq analyses also failed to demonstrate significant differences or trends in differentially expressed genes.

## Discussion

We describe 14 individuals harbouring biallelic *RNU2-2* variants presenting with a severe DEE phenotype. These variants clustered in the functionally important *RNU2-2* 5’ region, were ultra-rare and were associated with aberrant splicing. Taken together, we provide a refined delineation of the recessive *RNU2-2* DEE based on stringently defined cases with ultra-rare, region-specific biallelic *RNU2-2* variants and a highly concordant clinical presentation.

The individuals in our cohort had a significantly similar phenotypic signature when compared against the broader cohort of >700 individuals with epilepsy who have undergone WGS at CMMS. Performing phenotypic analyses against other individuals with complex epilepsies is advantageous as it directly reflects the specialised clinical diagnostic workflow and backdrop against which individuals with a suspected genetic epilepsy are assessed. Further, significant phenotypic differences are more likely to indicate a more specific shared phenotypic signature and a shared molecular aetiology than analyses where individuals’ phenotypic signatures are compared against broader disease cohorts. Since we had to exclude HPO terms identified through deep phenotyping to prevent bias our cohort likely share even greater phenotypic homogeneity than could be captured in these analyses. All but two individuals had seizure onset at or before the age of two years, and all individuals had refractory seizures and multiple seizure types, with tonic (100%), infantile/epileptic spasms (∼80%), bilateral tonic-clonic (∼60%), generalised atonic (∼50%) and generalised myoclonic (∼50%) seizures being especially prevalent. Severe to profound intellectual disability was seen in all individuals, with developmental delay being apparent within the first year of life for most cases. Developmental regression was observed in all individuals, with many losing skills during periods of increased seizure severity. Involuntary movements were a marked feature of our cohort presenting as choreoathetosis, dystonia or non-epileptic myoclonus. Further, many individuals lacked any voluntary movement and were unable to sit unsupported or walk independently.

When comparing the dominant and recessive *RNU2-2* disorders, there is considerable phenotypic overlap as well as distinctive traits unique to each condition. Both disorders are characterised by refractory, early childhood onset epilepsy with severe to profound intellectual disability and a high prevalence of comorbid microcephaly, obstructive sleep apnoea/sleep problems and hypotonia (/abnormal tone). The recessive disorder presents with a more severe motor impairment and a higher proportion of individuals with an abnormal brain MRI than the dominant condition. Our cohort had an earlier median seizure onset age of 10.5 months compared with 21 months in the dominant cases^5^. There is overlap with the seizure types across both conditions however our cohort had a higher proportion of individuals with epileptic spasms, tonic, atonic, myoclonic and absence seizures and individuals with LGS (like) and/or infantile epileptic spasms syndrome diagnoses^3–5^.

All the variants described in our cohort, with the exception of n.14C>G, were recurrent in our cohort and/or at the same positions as other biallelic *RNU2-2* variants reported recently in other studies (preprints) which have independently identified the same recessive *RNU2-2* disorder^13–15^. The prevalence of the biallelic *RNU2-2* disorder across suspected genetic epilepsies analysed at CMMS was 0.65% indicating that this is a relatively common “rare” disease.

A complex interplay of conformational changes and associations of the snRNPs and splicing factors is required for the execution of correct splicing. U2 snRNA is a core component in all stages of splicing from spliceosome assembly, activation, catalysis to disassembly. U2 snRNP, together with snRNPs U1, U4, U5 and U6, comprise the major spliceosome and execute splicing through interactions at key regions in each snRNP.

There are four highly conserved regions in U2 snRNA: stem loop I, the branch point recognition sequence (BPRS), stem loop II region and the Sm binding site, all of which are established as functionally important in splicing. Stem loop I is the region found most proximal to the 5’ end. Unwinding of stem loop I enables base pairing with U6 to form the U2/U6 helix Ia and Ib. U2/U6 helix Ia and Ib are important for splicing catalysis^27^, with mutations in yeast and humans being shown to hinder^28^ or abolish splicing^29^, respectively. Multiple individuals in our cohort harboured variants in stem loop I including n.23A>G and n.25G>T which fall within U2/U6 helix Ia and Ib^30^ thus may interrupt splicing catalysis. Variants earlier in stem loop I at nucleotides 3-14 abolished splicing^31^ in HEK 239 cells, thus the n.14C>G may also impair splicing. The recurrent pathogenic dominant *RNU2-2* n.4G>A variant is also located early in stem loop I^3–5^.

The branch point recognition site (BPRS) located downstream of stem loop I forms a branch point-interacting stem loop (BSL) early in spliceosome assembly^32^, which may act to temporally prevent the BPRS from binding with the pre-mRNA branch sequence (BS) until necessary. Displacement of splicing factor HTATSF1 disrupts BSL stability, enabling the pre-mRNA BS to associate with U2 snRNA through the BPRS and form the branch helix^33^. The U2/BS helix formation causes the BS adenosine to bulge out, making it a target for nucleophilic attack which is essential for the first catalytic step in splicing and formation of the B* complex^27^. Three recurrent variants, n.25G>T, n.44T>A and n.45C>T, identified in the current cohort, lie in the BSL at positions which base pair to create loop forming bonds, as well as in the BPRS, thus may disrupt splicing catalysis.

Stem loop II commences just downstream of the BPRS/BSL where *RNU2-2* variants n.52G>A, n.61C>G and n.61C>A identified in this study are located. Stem loop IIa is essential for cell viability^34^ and may be involved in the docking of the U2/BS helix into the active site of the spliceosome^27^.

During snRNP biogenesis, Sm proteins associate with pre-snRNAs at the Sm site and form the Sm ring, enabling snRNP maturation and stability^35^. The Sm binding site also influences splicing in an indirect manner through reorienting the snRNP relative to the spliceosome active site^27^. Sm binding site variants in *RNU4ATAC* cause a recessive minor spliceosome disorder through reducing cellular U4atac snRNP levels and destabilising U4atac snRNA^36^. The variant n.104T>G and the recurrent deletion (n.107_118del) in *RNU2-2*, identified in individuals from this cohort, lie within and just downstream of the Sm binding site, respectively. These variants may therefore disrupt U2 snRNA maturation and stability.

Over 95% of multi-exon genes undergo alternative splicing^37^ in a temporal and tissue specific manner, thus even mild splicing interruptions may have widespread effects. We explored RNA sequencing analyses across both blood and fibroblasts to better understand the pathophysiological mechanisms driving the recessive *RNU2-2* disorder. *RNU2-2* has high expression in the developing brain^4^ thus likely plays a prominent role in brain development. Fibroblasts exhibit significantly higher expression of NDD and epilepsy genes than blood^38^ and are minimally invasive to obtain. This is the first study to explore RNA sequencing data in any *RNU2-*2 or *RNU4-2* disorder in fibroblasts. Our analyses revealed that the biallelic *RNU2-2* disorder leads to aberrant splicing across all splicing events, with the most pronounced differences observed for mutually exclusive exon and alternate 3’ splice site events in fibroblasts. Previous RNA sequencing studies into *RNU2-2*^3^ and *RNU4-2*^6^ in blood have failed to find significant splicing differences, with the exception of a study by Nava *et al.*^8^ where they found pronounced differences at 5’ splice sites in individuals with dominant *RNU4-2* variants. We validated this finding in our *RNU4-2* cohort where we also observed 5’ splice site to be the most distinct event type. Interestingly, we also observed significant splicing differences across all splicing event types in both blood and fibroblasts suggesting that our analyses had greater detection sensitivity. The tissue analysed is one factor which improves sensitivity for detecting splicing differences in both *RNU2-2* and *RNU4-2* disorders. However, the additional differences detected in *RNU4-2* in the blood in the current study are more difficult to explain. A strength of the current study is that all RNA sequencing samples, preparation, sequencing and analyses were performed using the same protocol, sample preparation and sequencing facility which may have improved sensitivity to detect differences. Overall, these analyses confirm that the biallelic *RNU2-2* disorder leads to aberrant splicing which is most apparent in fibroblasts, reflective of the prominently neurological phenotype seen in these patients. Contrastingly, the dominant *RNU4-2* disorder, which exhibits a broader multisystem phenotype, displays more widespread, less tissue specific splicing abnormalities. Our findings are in agreement with other recessive minor spliceosomopathies which are also characterised by significant aberrant splicing events^39^, with a recent study demonstrating that a transcriptome first approach is sufficient to identify individuals with previously unknown rare, biallelic variants in minor spliceosome snRNA genes^10^.

In summary our study provides support for a distinct, severe DEE phenotype associated with ultra-rare, 5’ clustering biallelic *RNU2-2* variants. Our segregation and RNA sequencing analyses provide further support for the pathogenicity of these variants which segregated with the disease phenotype and implicates abnormal splicing, in particular mutually exclusive exon and alternative 3’ splice site events, as a pathophysiological mechanism. Novel *RNU2-2* variants identified in the future will likely require a combination of deep phenotyping, segregation and RNA sequencing analyses for accurate interpretation of pathogenicity.

## Acknowledgements

We would like to thank the study participants and their families for their invaluable contribution to this research. We thank the Clinical Genomics Stockholm facility at the Science for Life Laboratory for their expertise and support in sequencing analyses. We acknowledge the contributions of the UDN Sweden network to case ascertainment and deep phenotyping, including multidisciplinary clinical evaluations and data integration within the GMCK-RD framework. Several authors in this publication are members of the European Reference Network of Rare Congenital Malformations and Rare Intellectual Disability ERN-ITHACA (EU Framework Partnership Agreement ID: 3HP-FPA ERN-01-2016/739516).

## Supplementary Material

Supplementary material is available at *Brain* online.

## Data availability

Further data that substantiate the findings of this study can be obtained from the corresponding author upon reasonable request. Public release of the data is restricted to maintain privacy and comply with ethical requirements.

## References

1. Henry OJ, Ygberg S, Barbaro M, et al. Clinical whole genome sequencing in pediatric epilepsy: Genetic and phenotypic spectrum of 733 individuals. Epilepsia. Published online April 4, 2025:epi.18403. doi:10.1111/epi.18403

2. Guerrini R, Conti V, Mantegazza M, Balestrini S, Galanopoulou AS, Benfenati F. Developmental and epileptic encephalopathies: from genetic heterogeneity to phenotypic continuum. Physiol Rev. 2023;103(1):433–513. doi:10.1152/physrev.00063.2021

3. Greene D, De Wispelaere K, Lees J, et al. Mutations in the small nuclear RNA gene RNU2-2 cause a severe neurodevelopmental disorder with prominent epilepsy. Nat Genet. Published online April 10, 2025. doi:10.1038/s41588-025-02159-5

4. Jackson A, Thaker N, Blakes A, et al. Analysis of R-loop forming regions identifies RNU2-2 and RNU5B-1 as neurodevelopmental disorder genes. Nat Genet. Published online May 29, 2025. doi:10.1038/s41588-025-02209-y

5. Chiu ATG, Bennett MF, Thiyagarajah H, et al. Pathogenic Variants in *RNU2*lJ*2* , a Non=:Jcoding Spliceosomal RNA , Cause a Distinctive Developmental and Epileptic Encephalopathy. Annals of Neurology. Published online November 19, 2025:ana.78071. doi:10.1002/ana.78071

6. Chen Y, Dawes R, Kim HC, et al. De novo variants in the RNU4-2 snRNA cause a frequent neurodevelopmental syndrome. Nature. 2024;632(8026):832–840. doi:10.1038/s41586-024-07773-7

7. Greene D, Thys C, Berry IR, et al. Mutations in the U4 snRNA gene RNU4-2 cause one of the most prevalent monogenic neurodevelopmental disorders. Nat Med. 2024;30(8):2165–2169. doi:10.1038/s41591-024-03085-5

8. Nava C, Cogne B, Santini A, et al. Dominant variants in major spliceosome U4 and U5 small nuclear RNA genes cause neurodevelopmental disorders through splicing disruption. Nat Genet. Published online May 16, 2025. doi:10.1038/s41588-025-02184-4

9. Wilkinson ME, Charenton C, Nagai K. RNA Splicing by the Spliceosome. Annu Rev Biochem. 2020;89:359–388. doi:10.1146/annurev-biochem-091719-064225

10. Arriaga TM, Mendez R, Ungar RA, et al. Transcriptome-wide outlier approach identifies individuals with minor spliceopathies. Am J Hum Genet. 2025;112(10):2458–2475. doi:10.1016/j.ajhg.2025.08.018

11. Stranneheim H, Lagerstedt-Robinson K, Magnusson M, et al. Integration of whole genome sequencing into a healthcare setting: high diagnostic rates across multiple clinical entities in 3219 rare disease patients. Genome Med. 2021;13(1):40. doi:10.1186/s13073-021-00855-5

12. Lindstrand A, Lagerstedt-Robinson K, Jemt A, et al. The Genomic Medicine Center Karolinska 10-year report on genome sequencing for rare diseases and a strategy for stepwise clinical implementation. In Review. Preprint posted online June 17, 2025. doi:10.21203/rs.3.rs-6790162/v1

13. Greene D, Mendez R, Lees J, et al. Biallelic variants in *RNU2-2* cause the most prevalent known recessive neurodevelopmental disorder. Genetic and Genomic Medicine. Preprint posted online August 29, 2025. doi:10.1101/2025.08.26.25334179

14. Jackson A, Blakes AJ, Wall E, et al. Biallelic variants in *RNU2-2* cause a remarkably frequent developmental epileptic encephalopathy. Genetic and Genomic Medicine. Preprint posted online September 4, 2025. doi:10.1101/2025.09.02.25334957

15. Leitão E, Santini A, Cogne B, et al. Systematic analysis of snRNA genes reveals frequent *RNU2-2* variants in dominant and recessive developmental and epileptic encephalopathies. Genetic and Genomic Medicine. Preprint posted online September 4, 2025. doi:10.1101/2025.09.02.25334923

16. Gargano MA, Matentzoglu N, Coleman B, et al. The Human Phenotype Ontology in 2024: phenotypes around the world. Nucleic Acids Res. 2024;52(D1):D1333–D1346. doi:10.1093/nar/gkad1005

17. Palmer EE, Cederroth H, Cederroth M, et al. Equity in action: The Diagnostic Working Group of The Undiagnosed Diseases Network International. NPJ Genom Med. 2024;9(1):37. doi:10.1038/s41525-024-00422-y

18. Magnusson M, Eisfeldt J, Nilsson D, et al. Loqusdb: added value of an observations database of local genomic variation. BMC Bioinformatics. 2020;21(1):273. doi:10.1186/s12859-020-03609-z

19. Greene D, De Wispelaere K, Lees J, et al. Mutations in the U2 snRNA gene RNU2-2P cause a severe neurodevelopmental disorder with prominent epilepsy. medRxiv. Published online September 4, 2024:2024.09.03.24312863. doi:10.1101/2024.09.03.24312863

20. Beniczky S, Trinka E, Wirrell E, et al. Updated classification of epileptic seizures: Position paper of the International League Against Epilepsy. Epilepsia. Published online April 23, 2025. doi:10.1111/epi.18338

21. Zuberi SM, Wirrell E, Yozawitz E, et al. ILAE classification and definition of epilepsy syndromes with onset in neonates and infants: Position statement by the ILAE Task Force on Nosology and Definitions. Epilepsia. Published online May 3, 2022. doi:10.1111/epi.17239

22. Specchio N, Wirrell EC, Scheffer IE, et al. International League Against Epilepsy classification and definition of epilepsy syndromes with onset in childhood: Position paper by the ILAE Task Force on Nosology and Definitions. Epilepsia. Published online May 3, 2022. doi:10.1111/epi.17241

23. Greene D, Richardson S, Turro E. ontologyX: a suite of R packages for working with ontological data. Bioinformatics. 2017;33(7):1104–1106. doi:10.1093/bioinformatics/btw763

24. Yépez VA, Mertes C, Müller MF, et al. Detection of aberrant gene expression events in RNA sequencing data. Nat Protoc. 2021;16(2):1276–1296. doi:10.1038/s41596-020-00462-5

25. Wang Y, Xie Z, Kutschera E, Adams JI, Kadash-Edmondson KE, Xing Y. rMATS-turbo: an efficient and flexible computational tool for alternative splicing analysis of large-scale RNA-seq data. Nat Protoc. 2024;19(4):1083–1104. doi:10.1038/s41596-023-00944-2

26. Mertes C, Scheller IF, Yépez VA, et al. Detection of aberrant splicing events in RNA-seq data using FRASER. Nat Commun. 2021;12(1):529. doi:10.1038/s41467-020-20573-7

27. Van Der Feltz C, Hoskins AA. Structural and functional modularity of the U2 snRNP in pre-mRNA splicing. Critical Reviews in Biochemistry and Molecular Biology. 2019;54(5):443–465. doi:10.1080/10409238.2019.1691497

28. Ryan DE, Abelson J. The conserved central domain of yeast U6 snRNA: importance of U2-U6 helix Ia in spliceosome assembly. RNA. 2002;8(8):997–1010. doi:10.1017/s1355838202025013

29. Sun JS, Manley JL. A novel U2-U6 snRNA structure is necessary for mammalian mRNA splicing. Genes Dev. 1995;9(7):843–854. doi:10.1101/gad.9.7.843

30. Anokhina M, Bessonov S, Miao Z, Westhof E, Hartmuth K, Lührmann R. RNA structure analysis of human spliceosomes reveals a compact 3D arrangement of snRNAs at the catalytic core. EMBO J. 2013;32(21):2804–2818. doi:10.1038/emboj.2013.198

31. Wu J, Manley JL. Multiple functional domains of human U2 small nuclear RNA: strengthening conserved stem I can block splicing. Mol Cell Biol. 1992;12(12):5464–5473. doi:10.1128/mcb.12.12.5464-5473.1992

32. Zhang Z, Will CL, Bertram K, et al. Molecular architecture of the human 17S U2 snRNP. Nature. 2020;583(7815):310–313. doi:10.1038/s41586-020-2344-3

33. Tholen J, Razew M, Weis F, Galej WP. Structural basis of branch site recognition by the human spliceosome. Science. 2022;375(6576):50–57. doi:10.1126/science.abm4245

34. Ares M, Igel AH. Lethal and temperature-sensitive mutations and their suppressors identify an essential structural element in U2 small nuclear RNA. Genes Dev. 1990;4(12A):2132–2145. doi:10.1101/gad.4.12a.2132

35. Pánek J, Roithová A, Radivojević N, et al. The SMN complex drives structural changes in human snRNAs to enable snRNP assembly. Nat Commun. 2023;14(1):6580. doi:10.1038/s41467-023-42324-0

36. Jafarifar F, Dietrich RC, Hiznay JM, Padgett RA. Biochemical defects in minor spliceosome function in the developmental disorder MOPD I. RNA. 2014;20(7):1078–1089. doi:10.1261/rna.045187.114

37. Pan Q, Shai O, Lee LJ, Frey BJ, Blencowe BJ. Deep surveying of alternative splicing complexity in the human transcriptome by high-throughput sequencing. Nat Genet. 2008;40(12):1413–1415. doi:10.1038/ng.259

38. Murdock DR, Dai H, Burrage LC, et al. Transcriptome-directed analysis for Mendelian disease diagnosis overcomes limitations of conventional genomic testing. J Clin Invest. 2021;131(1):e141500, 141500. doi:10.1172/JCI141500

39. Norppa AJ, Shcherbii MV, Frilander MJ. Connecting genotype and phenotype in minor spliceosome diseases. RNA. 2025;31(3):284–299. doi:10.1261/rna.080337.124

