## Supplementary material for "Phenotypic and transcriptomic characterisation of a novel biallelic *RNU2-2* developmental and epileptic encephalopathy"

### SUPPLEMENTARY DATA

#### Methods

##### HPO similarity analyses

HPO terms included in the phenotype similarity analyses included: seizure (HP:0001250), focal seizure (HP:0007359), generalised seizure (HP:0002197), infantile spasms (HP:0012469), neonatal seizure onset (HP:0003623), infantile seizure onset (HP:0003593), childhood seizure onset (HP:0011463), neurodevelopmental abnormality (HP:0012759), global developmental delay (HP:0001263), intellectual disability (HP:0001249), autistic behaviour (HP:0000729), movement abnormality (HP:0100022), abnormal muscle tone (HP:0003808), microcephaly (HP:0000252), brain imaging abnormality (HP:0410263), death in childhood (HP:0003819).

### Results

#### Ultra-rare biallelic RNU2-2 variants segregate with a DEE phenotype

Two individuals with incompatible phenotypes to the biallelic *RNU2-2* cohort also presented with biallelic *RNU2-2* variants including one rarer variant prior to n.61. One individual harboured the recurrent n.45C>T and n.148C>T, which is more common (73 heterozygotes gnomAD v4.1nonUKB) and in not recurrent in any of the reported studies. The second individual harboured variants n.54T>C and n.106G>C however neither variant is previously reported and this individual also harbours a causative pathogenic variant in a different gene. Thus, these variant combinations are considered variants of uncertain significance (VUS) and appear unlikely to contribute to the severe biallelic DEE disorder.

#### Supplementary figures

**Figure 1**. **rMATS-turbo splicing analyses across all events in blood and fibroblasts in *RNU2-2* individuals compared with controls. A)** 3’ splice sites in blood. **B)** 5’ splice sites in blood. **C)** Mutually exclusive exons in blood. **D)** Retained introns in blood. **E)** Skipped exons in blood. **F)** Retained introns in fibroblasts. **G)** 5’ splice sites in fibroblasts. **H)** Skipped exons in fibroblasts.

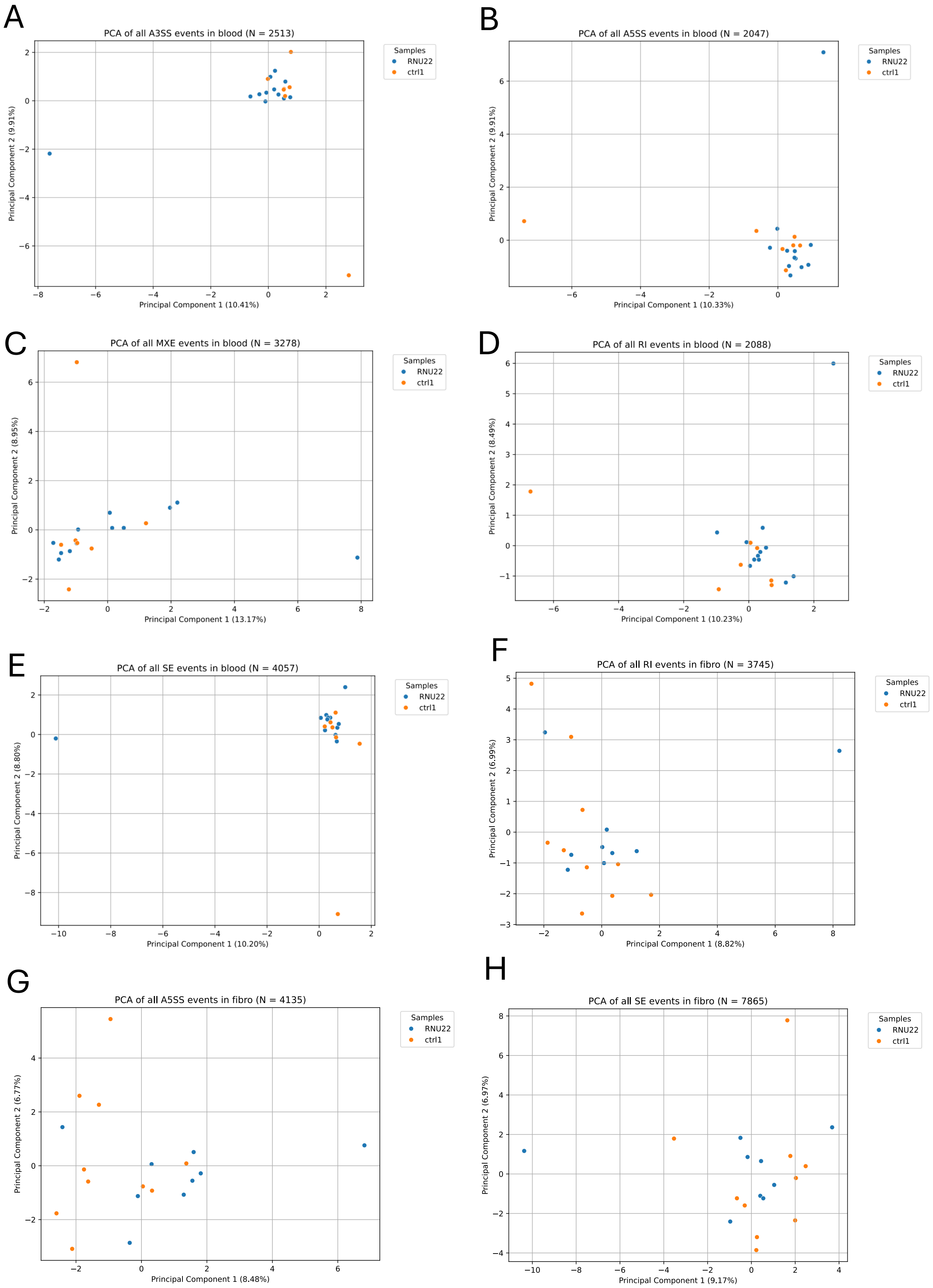

**Figure 2. rMATS-turbo splicing analyses across significant events in blood and fibroblasts *RNU2-2* individuals compared with controls. A)** Mutually exclusive exons in blood. **B)** 3' splice sites in blood. **C)** 5' splice sites in blood. **D)** Retained introns in blood**. sE)** Skipped exons from blood. **F)** 5' splice sites in fibroblasts. **G)** Retained introns in fibroblasts. **F)** Skipped exons in fibroblasts.

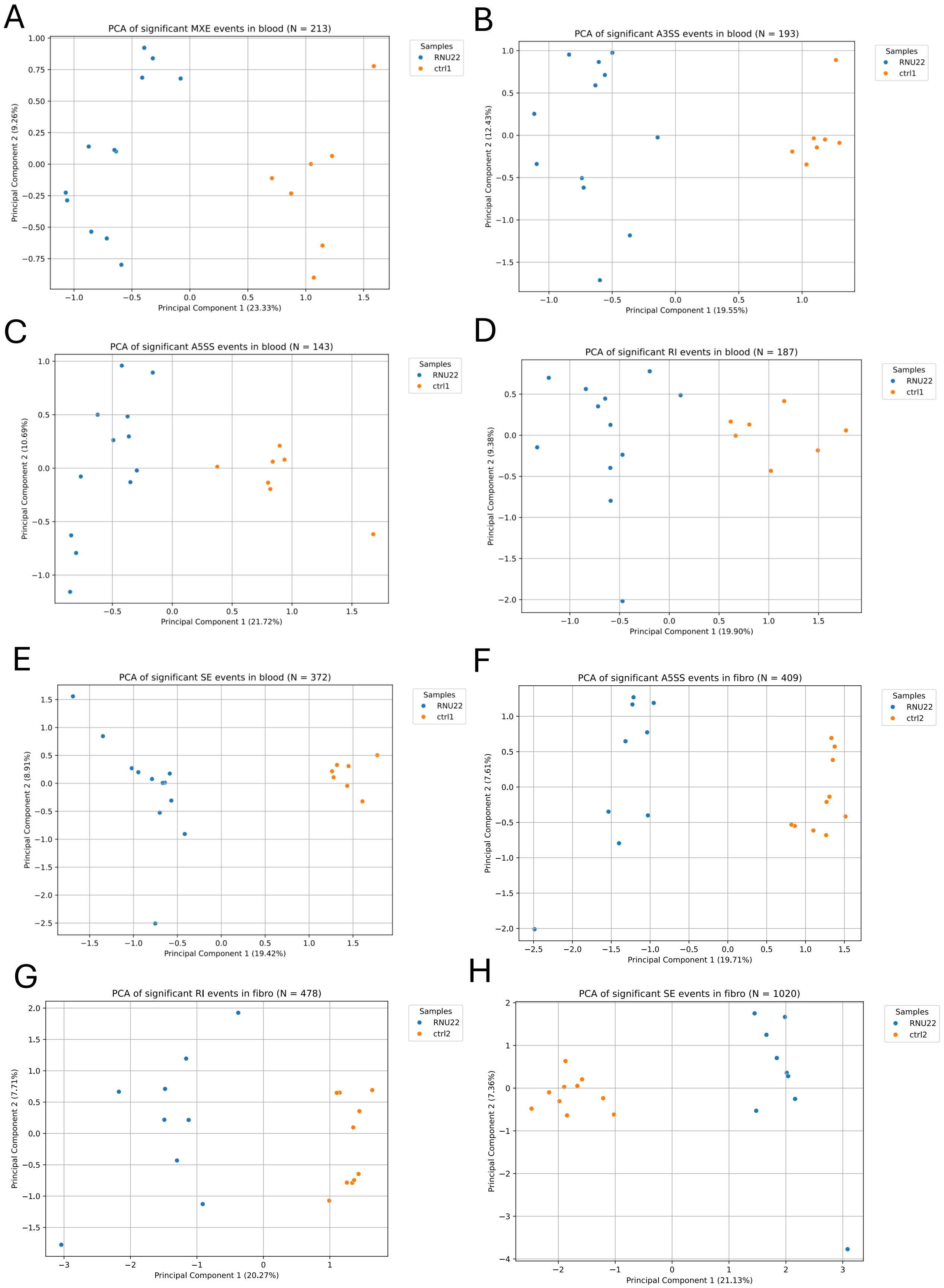

**Figure 3. rMATS-turbo splicing analyses across all events in blood and fibroblasts in *RNU4-2* individuals compared with controls. A)** 5' splice sites in blood. **B)** 5' splice sites in fibroblasts. **C)** 3' splice sites in fibroblasts. **D)** Mutually exclusive exons in fibroblasts. **E)** Retained introns in fibroblasts. **F)** Skipped exons in fibroblasts.

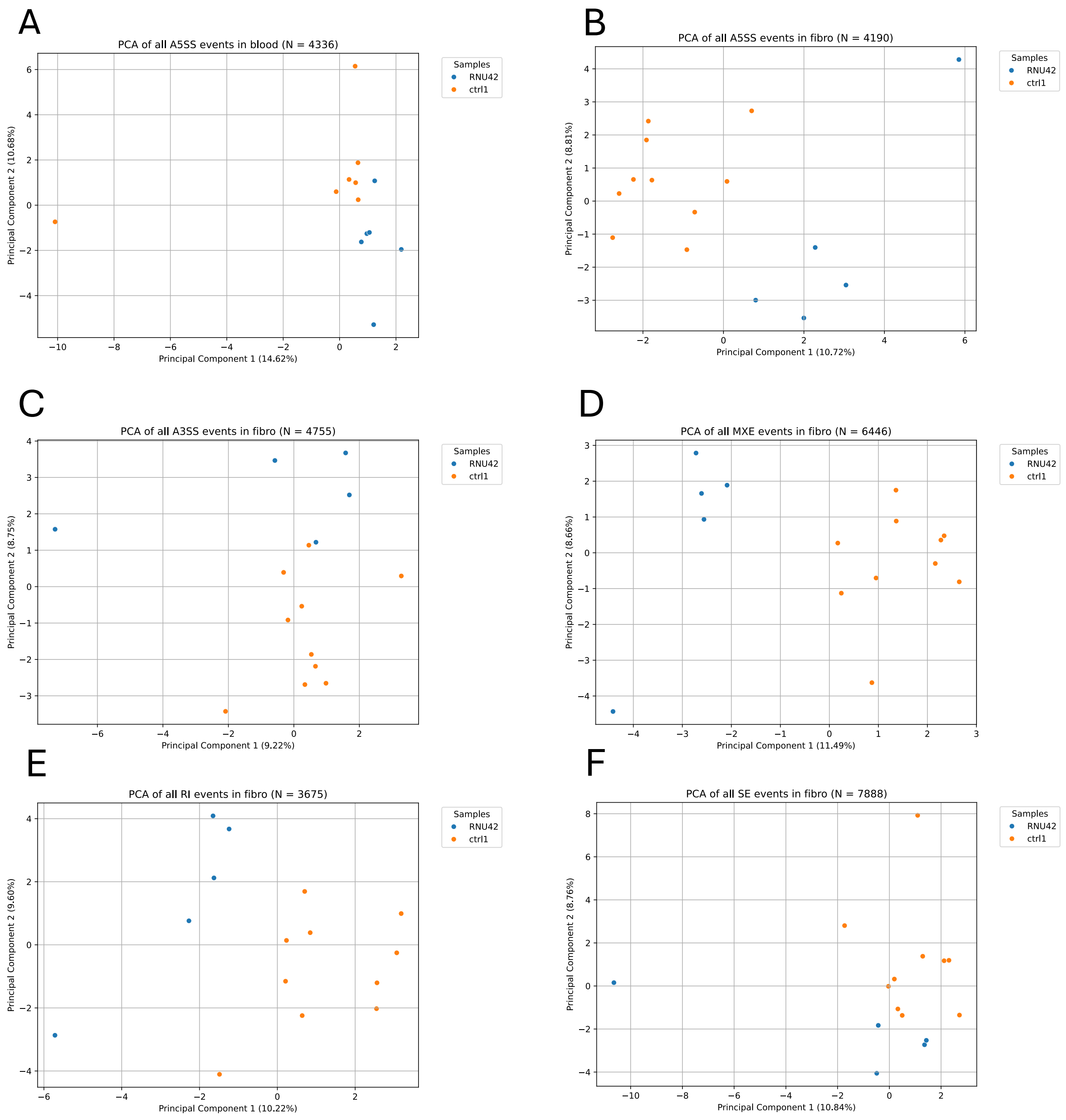

**Figure 4. rMATS-turbo splicing analyses across significant events in blood and fibroblasts in *RNU4-2* individuals compared with controls. A) Mutually exclusive exons in fibroblasts. B)** Mutually exclusive exons in blood.**C)** 3’ splice sites in fibroblasts. **D)** 3’ splice sites in blood. **E)** 5’ splice sites in fibroblasts. **F)** 5’ splice sites in blood. **G)** Retained introns in fibroblasts. **H)** Retained introns in blood. **I)** Skipped exons in fibroblasts. **J)** Skipped exons in blood.

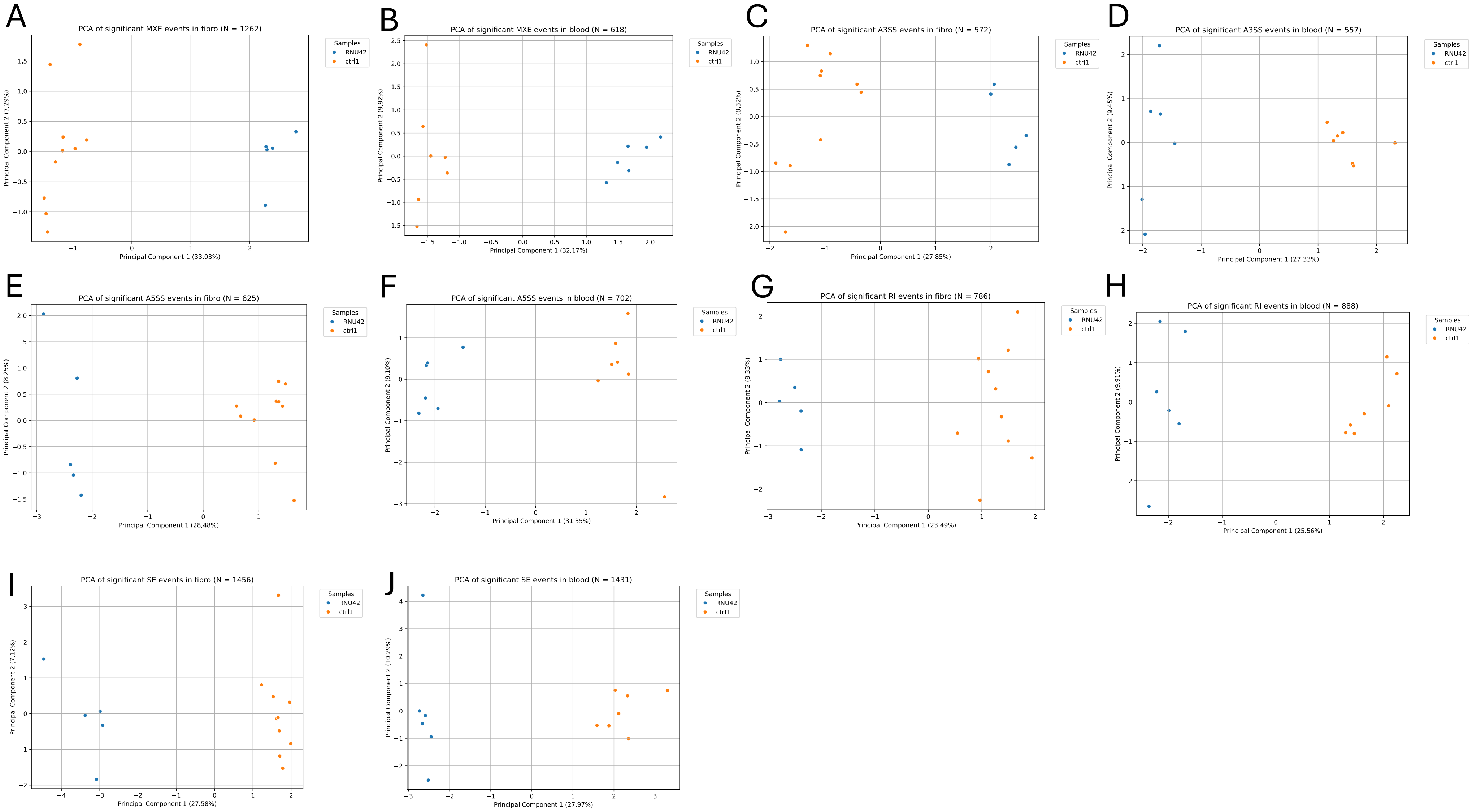

**Figure 5. DROP RNA sequencing analyses in fibroblasts in *RNU2-2* and *RNU4-2* individuals compared with controls.** A red star denotes a significant ($p \leq 0.05$) difference from controls. **A)** Aberrant expression events from OUTRIDER across all genes. **B)** Aberrant splicing events from FRASER across all genes. **C)** Aberrant expression events from OUTRIDER from the CMMS version 22 epilepsy panel genes. **D)** Aberrant splicing events from FRASER from the CMMS version 22 epilepsy panel genes.

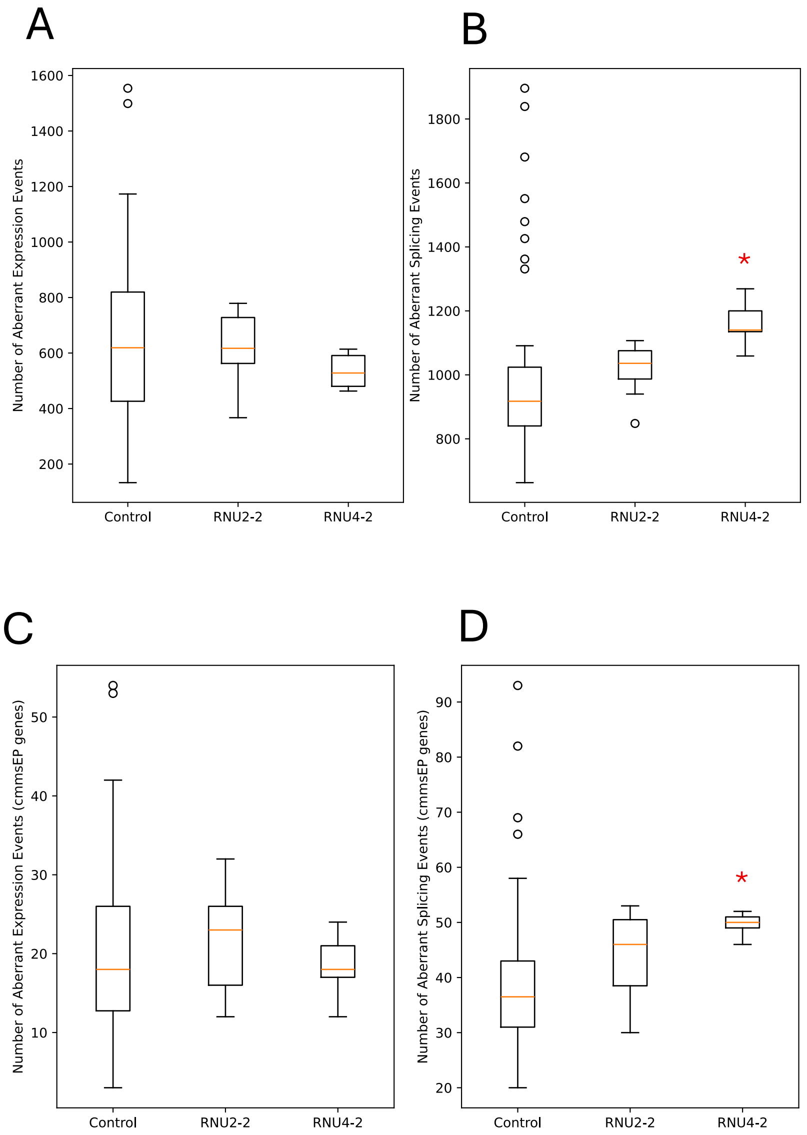

**Figure 6. Sashimi plots from rMATS analysis showing isoform shifts in *MAP4K4* and *AKNA* in *RNU4-2* individuals compared with controls in blood.** Isoform shifts seen in *MAP4K4* and *ANKA* are similar to findings presented by Nava *et al.*[1] **A)** *MAP4K4 RNU4-2* isoform shift 42.4% compared with 18.3% in controls. **B)** *AKNA RNU4-2* isoform shift 14.4% compared with 0.3% in controls.

A

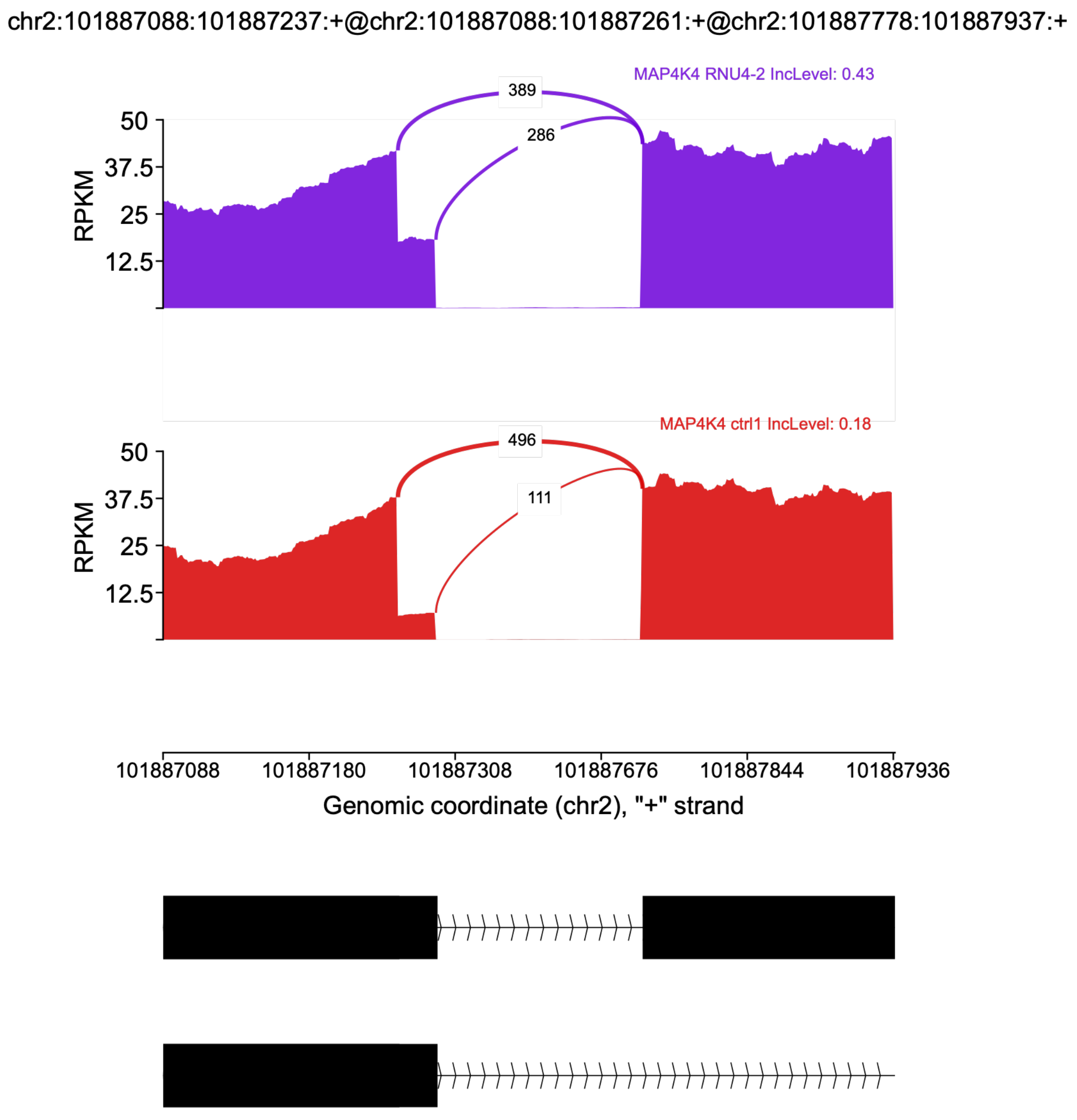

B

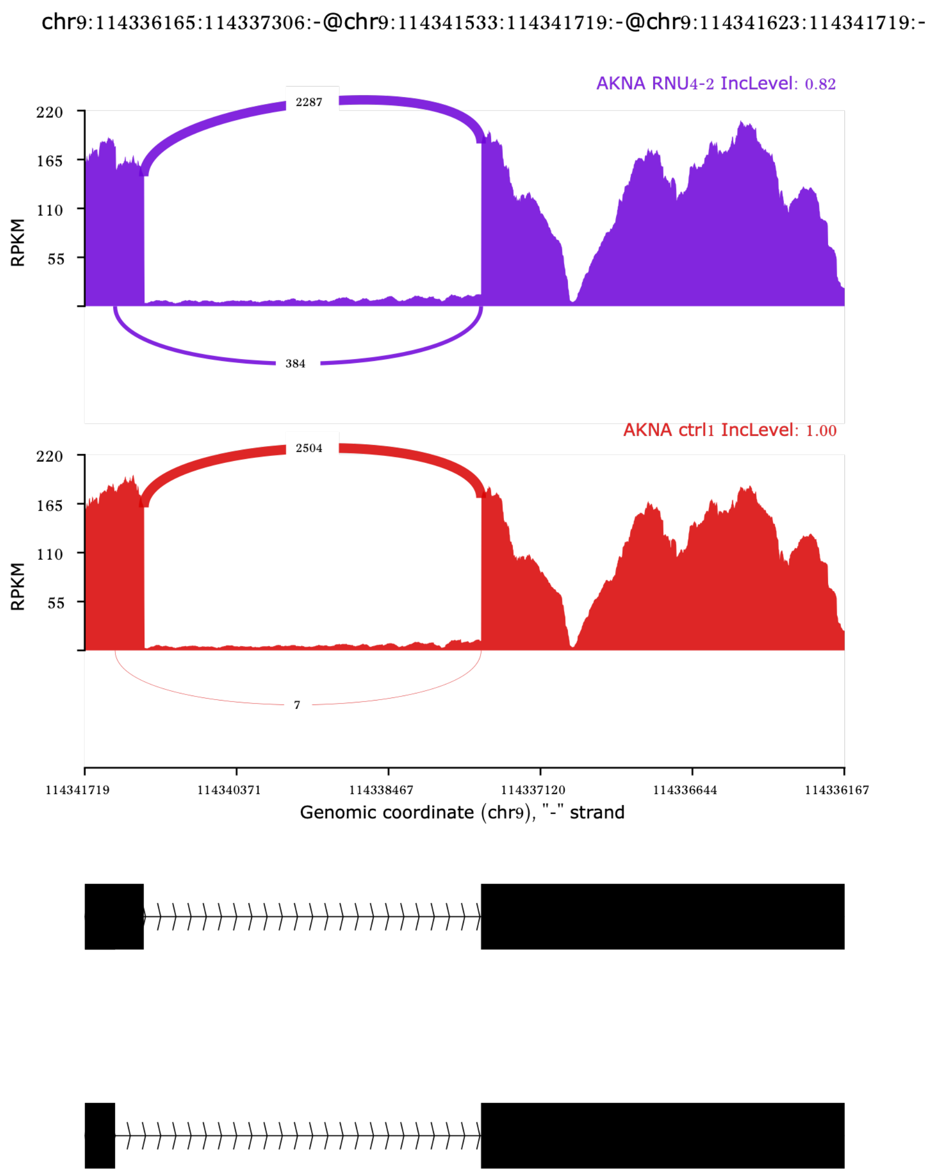
chr9:114336165:114337306:-@chr9:114341533:114341719:-@chr9:114341623:114341719:-
AKNA RNU4-2 IncLevel: 0.82
2287
384
RPKM
220
165
110
55
AKNA ctrl1 IncLevel: 1.00
2504
7
114341719
114340371
114338467
114337120
114336644
114336167
Genomic coordinate (chr9), "-" strand

### Supplementary references

1. Nava C, Cogne B, Santini A, et al. Dominant variants in major spliceosome U4 and U5 small nuclear RNA genes cause neurodevelopmental disorders through splicing disruption. *Nat Genet*. Published online May 16, 2025. doi:10.1038/s41588-025-02184-4
